# The management of Moderate Acute Malnutrition (MAM) in children aged 6-59 months: A systematic review and meta-analysis

**DOI:** 10.1101/2021.01.16.21249861

**Authors:** I Gluning, M Kerac, J Bailey, A Bander, C Opondo

**Affiliations:** Department of Population Health, London School of Hygiene and Tropical Medicine, London, UK; Brighton and Sussex University Hospitals Trust, Brighton, UK; Centre for MARCH (Maternal, Reproductive, Adolescent & Child Health Centre), London School of Hygiene and Tropical Medicine, London, UK; International Rescue Committee, New York, NY, USA; Department of Medical Statistics, London School of Hygiene and Tropical Medicine, London, UK; National Perinatal Epidemiology Unit, University of Oxford, Oxford, UK

## Abstract

**Background:** Malnutrition is a leading cause of morbidity and mortality in children aged under five years, especially in low- and middle-income countries (LMICs). Although severe acute malnutrition (SAM) is considered the most serious form of malnutrition, moderate acute malnutrition (MAM) affects greater numbers globally and, unlike SAM, guidelines lack a robust evidence-base. This systematic review and meta-analysis assessed the evidence for lipid-based nutrient supplements (LNS), fortified-blended-flours (FBF) and nutrition counselling, in the treatment of MAM.

**Methods:** Five databases were systematically searched for studies conducted in LMICs that compared the effectiveness of food-based products versus any comparator group in promoting recovery from MAM in children aged 6-59 months. Where appropriate, pooled estimates of effect were estimated using random-effects meta-analyses.

**Results:** A total of thirteen trials were identified for inclusion. All used active controls rather than ‘standard care’, which is often minimal in most settings. There was evidence of increased probability of recovery (as assessed by gaining normal weight-for-height and/or MUAC) among children treated with LNS compared to children treated with FBF (RR 1·05, 95%CI 1·01-1·09, p=0·009). Treatment with an LNS was also associated with a lower risk of persistent MAM at the end of treatment compared with a FBF (RR 0·82, 95%CI 0·71-0·95, p=0·007).

**Conclusion:** Based on a relatively small number of studies mainly from Africa, LNS are superior to FBF in improving anthropometric recovery from MAM. The true benefit of MAM treatment may be underestimated due to all studies using active controls rather than usual care which is minimal.

More high-quality evidence is needed to evaluate nutrition education/counselling alone as a MAM intervention. Studies should also assess a wider range of outcomes including body composition, morbidity and development – not weight-gain alone.

## Introduction

Malnutrition is a leading cause of morbidity and mortality in children aged under five, especially in low- and middle-income countries (LMIC)^1^. Acute malnutrition (AM) comprises wasting and/or nutritional oedema. Wasting is defined by low weight-for-length/height z-score (WLZ/WHZ) and/or low mid-upper arm circumference (MUAC). AM is often subdivided into severe acute malnutrition (SAM) and moderate acute malnutrition (MAM). In 2019, of the 47 million children under five years of age who were acutely malnourished, 32·7 million suffered from MAM^1^.

To date, much focus has been on treating children with SAM since individual case-fatality is higher. However, MAM also matters: it affects greater numbers of children globally; children with MAM have a three-fold increased risk of mortality compared to those without MAM; children with MAM are at-risk of deteriorating to SAM and undergo poorer physical and cognitive development compared to their well-nourished counterparts^1,2,3^

MAM treatment is context-specific and commonly involves one of two options: 1) improving the adequacy of the home diet through nutrition education/counselling^4^, or 2) supplementary feeding with an energy-dense product. The latter may be necessary in settings with food insecurity or where dietary diversity is poor, and involve targeted supplementary feeding programmes (SFPs)^5^. In recent years, treatment of MAM has predominantly focused on the use of fortified blended flours (FBF) and lipid-based nutrient supplements (LNS) as standard care^5,6^. A key barrier to scale-up is the lack of World Health Organization (WHO) guidelines on MAM. This review aims to inform an upcoming 2021 WHO guideline review where MAM (moderate wasting) is one of four key topics being examined^7^.

## Methods

We followed the PRISMA (Preferred Reporting-Items for Systematic Reviews and Meta-Analyses) reporting framework^8^.

### Inclusion Criteria

Eligibility and inclusion into the review were based on the following PICO (Population, Intervention, Comparator, and Outcome) framework:

- Children aged 6-59 months diagnosed with MAM and living in a LMIC. MAM was defined as having a WHZ <-2 and ≥-3 based on *WHO 2006 Growth Standards*^9^, and/or a MUAC <12·5cm and ≥11·5cm, without bipedal oedema.
- A supplementary food product used for the treatment of MAM. This includes: FBFs such as corn-soy blend (CSB), LNSs or ready-to-use therapeutic/supplementary food (RUTF/RUSF), or any complementary food supplement to be consumed in addition to the home diet.
- A comparative treatment group containing participants with MAM, who are receiving either: no specific intervention (‘usual care’ control, since MAM is not routinely identified or treated in all settings); active control, which includes an alternative food/supplement, nutrition education/counselling, any intervention/combination of interventions used to treat MAM.
- The primary outcome of interest was recovery from MAM, defined as having a WHZ >-2, and/or a MUAC >12·5cm, without, bipedal oedema, and based on *WHO 2006 Growth Standards*. Secondary outcomes of interest were; persistent MAM, progression to SAM, death, defaulting, and any adverse-effects to treatment including body composition.

Only published studies with a control/comparator group were included. For generalizability and inter-study comparability, we only included studies from 2006 to present (2020): 2006 marking the development of the *WHO 2006 Growth Standards* for defining MAM and SAM, taking over the previously used *NCHS References*.

### Search Methods

The following databases were searched by two authors independently, with final search completed on the 29^th^ October 2020: EMBASE (Ovid), MEDLINE (Ovid), CINAHL (EBSCO), Global Health and ClinicalTrials.gov.

Searches were limited to human-only studies, published between the years 2006 and 2020, and written in English language. A detailed search strategy is in *Appendix 1*.

The reference lists of identified studies were also checked for papers that met inclusion criteria.

### Data Collection

Studies identified in the search were initially screened by title and abstract to determine if they met eligibility criteria. They were then screened from their full text. Reasons for exclusion were documented.

Data was collected using a data collection form (*Table 1*) generated from the ‘checklist of items to consider in data collection or data extraction,’ as part of the Cochrane Methods handbook^10^.

**Table 1:**
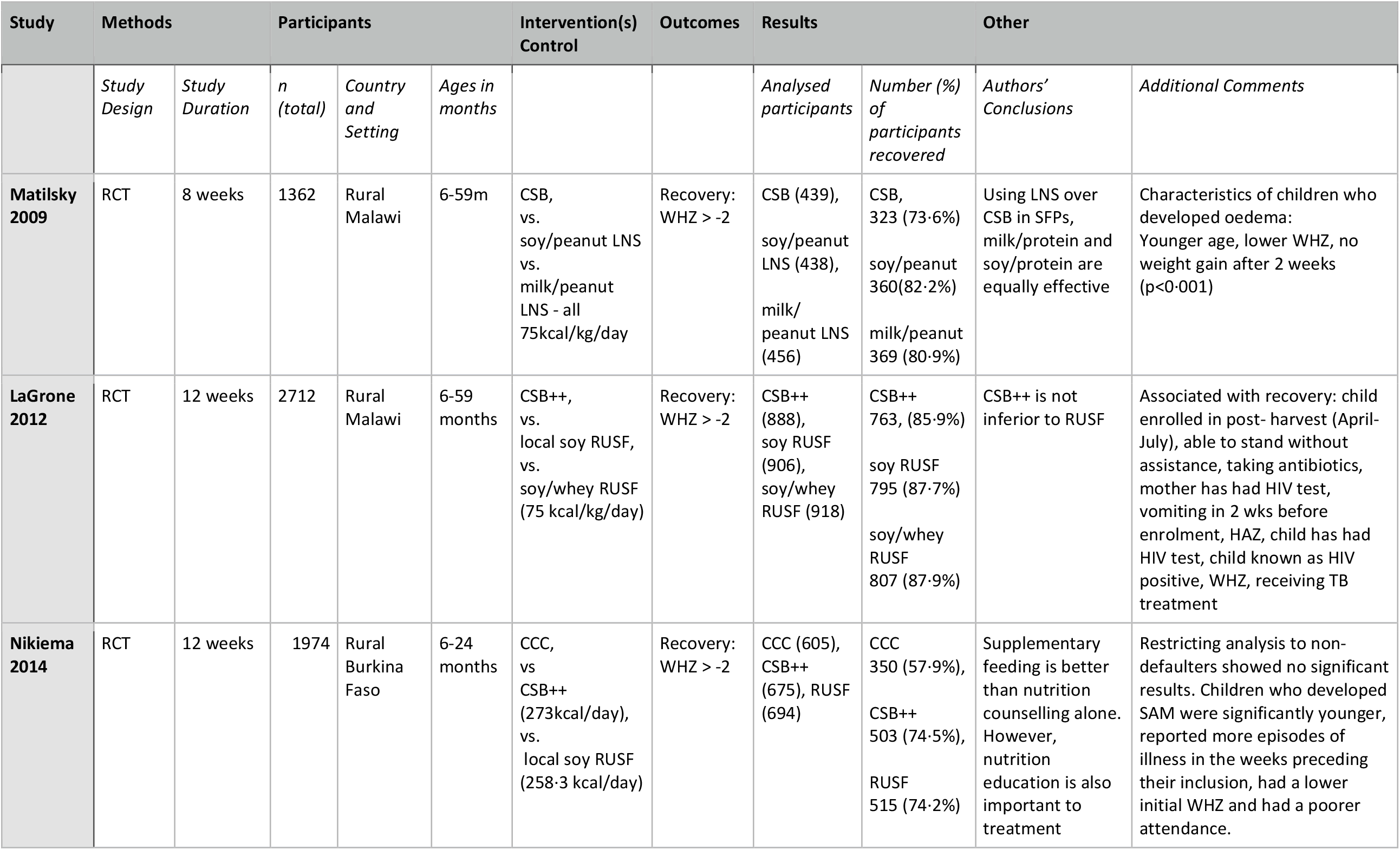
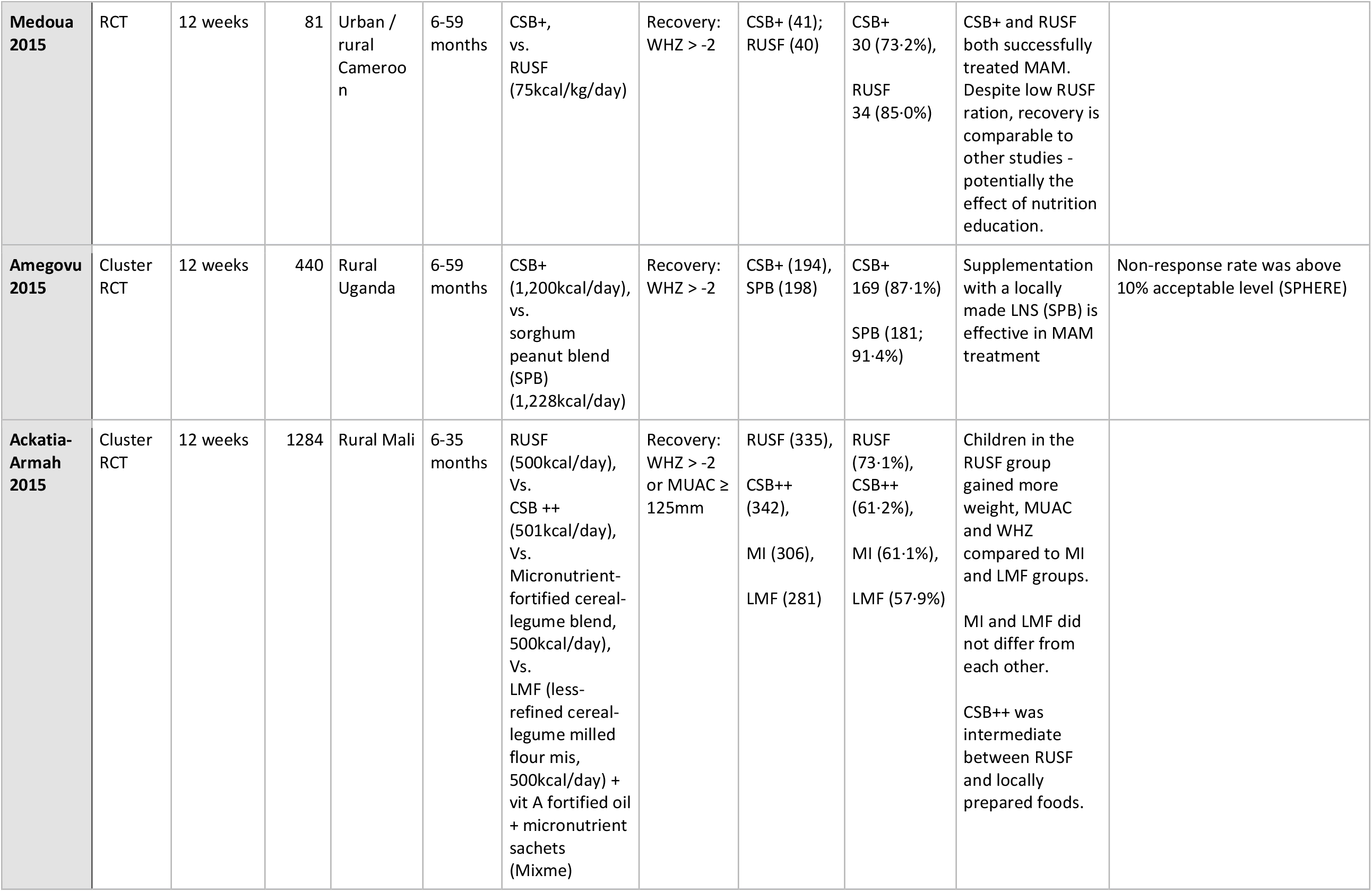
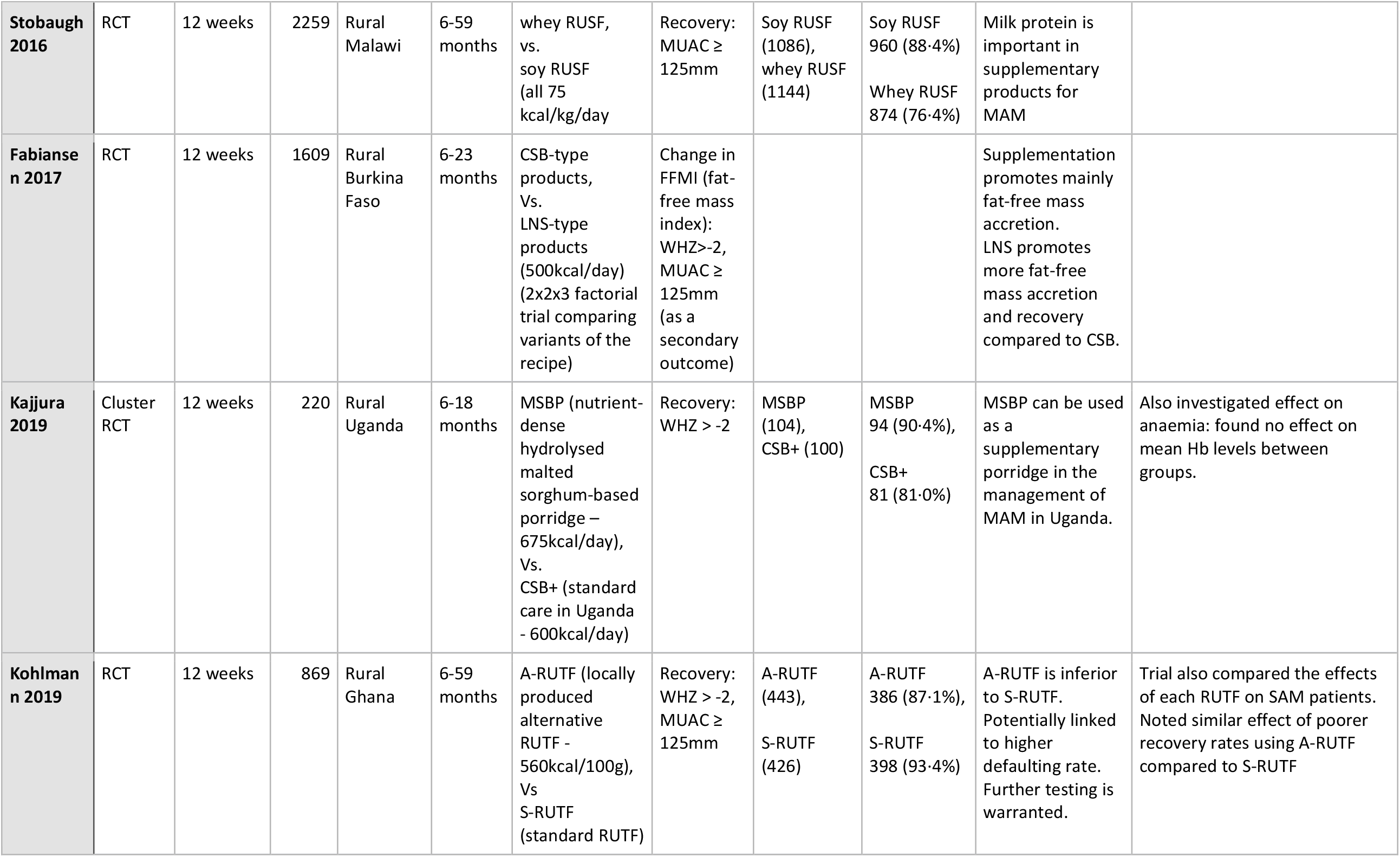
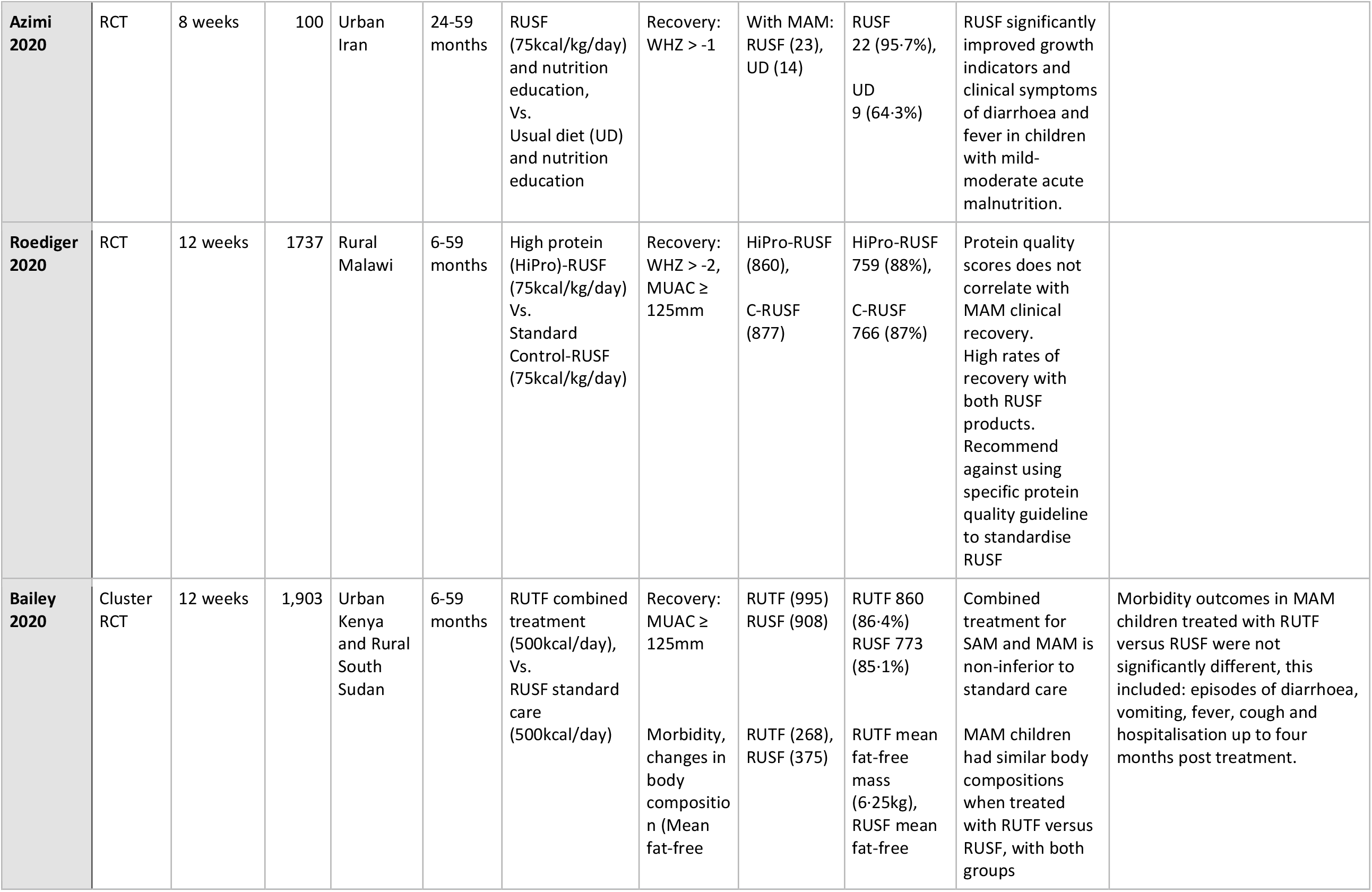
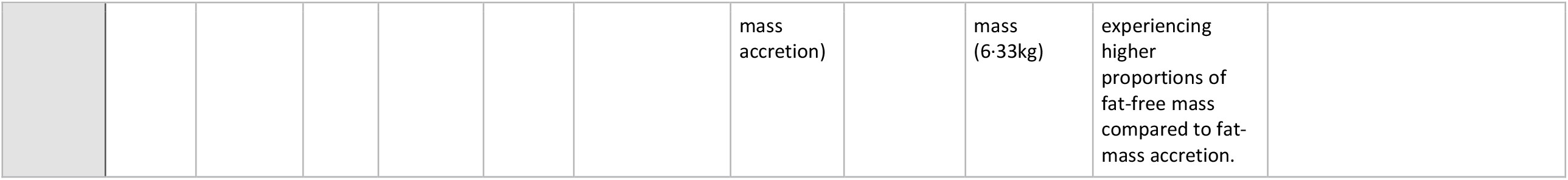
Data collection table of included studies Study characteristics for the thirteen studies included in this systematic review and meta-analysis in order of publication year. Important abbreviations: CCC, child-centred counselling; CSB, corn-soy blend; HAZ, height-for-age z-score; LNS, lipid-based nutrient supplement; MAM, moderate acute malnutrition; RCT, randomized controlled trial; RUSF, ready to use supplementary food; RUTF, ready to use therapeutic food; SAM, severe acute malnutrition; SFP, supplementary feeding programme; WHZ, weight-for-height z-score

**Table 1:** Risk of bias assessment for the thirteen included studies in order of publication year.

| **Study** | *Random Sequence Generation* | *Allocation Concealment* | *Blinding of Participants and Personnel* | *Blinding of Outcome Assessment* | *Incomplete Outcome Data* | *Selective Reporting* | *Other Sources of Bias* | **Overall Assessment** |
| --- | --- | --- | --- | --- | --- | --- | --- | --- |
| **Matilsky 2009** | Unclear Risk | Low Risk | Unclear Risk | Low Risk | Low Risk | Unclear Risk | Low Risk | **Good** |
| **LaGrone 2012** | Low Risk | Low Risk | Unclear Risk | Low Risk | Low Risk | Unclear Risk | Low Risk | **Good** |
| **Nikiema 2014** | Unclear Risk | Unclear Risk | Unclear Risk | Unclear Risk | High Risk | Unclear Risk | Low Risk | **Poor** |
| **Stobaugh 2016** | Unclear Risk | Low Risk | Low Risk | Low Risk | Low Risk | Unclear Risk | Low Risk | **Good** |
| **Medoua 2015** | Low Risk | Low Risk | Unclear Risk | Low Risk | Low Risk | Unclear Risk | Low Risk | **Good** |
| **Amegovu 2015** | High Risk | Low Risk | Unclear Risk | High Risk | Low Risk | Unclear Risk | Low Risk | **Poor** |
| **Ackatia-A 2015** | Unclear Risk | Unclear Risk | High Risk | Unclear Risk | Low Risk | Unclear Risk | Low Risk | **Poor** |
| **Fabiansen 2017** | Low Risk | Unclear Risk | Unclear Risk | Low Risk | Low Risk | Unclear Risk | Low Risk | **Good** |
| **Kajjura 2019** | Low Risk | Low Risk | Unclear Risk | Low Risk | Low Risk | Unclear Risk | Low Risk | **Good** |
| **Kohlmann 2019** | Low Risk | Low Risk | Low Risk | Low Risk | Unclear Risk | Unclear Risk | Low Risk | **Good** |
| **Azimi 2020** | Low Risk | Unclear Risk | High Risk | Unclear Risk | Low Risk | Low Risk | Low Risk | **Good** |
| **Roediger 2020** | Low Risk | Low Risk | Low Risk | Low Risk | Low Risk | Unclear Risk | Low Risk | **Good** |
| **Bailey 2020** | Low Risk | Low Risk | High Risk | Low Risk | Low Risk | Low Risk | Low Risk | **Good** |

Additional information was sought on the differences in characteristics of children who recovered from MAM, versus those who did not recover/progressed to SAM/defaulted/died.

### Risk of Bias

Risk of bias was determined using the *Cochrane Collaboration’s Tool for Assessing Risk of Bias*^11^, which considers the following: 1) random sequence generation, 2) allocation concealment, 3) blinding of participants and personnel, 4) blinding of outcome assessment, 5) incomplete outcome data, and 6) selective reporting.

For each criterion, a judgement was made as to whether the trial was at a ‘low-risk,’ a ‘high-risk,’ or an ‘unclear-risk’ of bias (*Table 2)*.

### Study Protocol

We pre-registered the study at www.crd.york.ac.uk/prospero/display_record.php?RecordID=68513.

### Data Analysis

For our primary and secondary outcomes we used STATA v·14^12^ to perform random-effects meta-analyses to estimate pooled risk ratios (RR) for the outcomes. Random-effects meta-analyses were chosen to incorporate the expected random variation in the effect of each intervention across the studies into the pooled estimates.

We also used descriptive synthesis to summarise findings about characteristics of children who recovered from MAM versus those who did not.

## Results

Our search (*Appendix 1)* generated 1,968 references after removing 309 duplicates. Thirteen papers were eligible for inclusion in the systematic review and meta-analysis: *Figure 1*. Reasons for exclusion of papers included: unpublished results; using NCHS references for defining MAM; using an alternative measurement of MAM (other than WHZ or MUAC); using a different definition for recovery from MAM (i.e. WHZ > −1), and/or investigating MAM prevention rather than MAM treatment (*Appendix 2)*.

**Figure 1:**
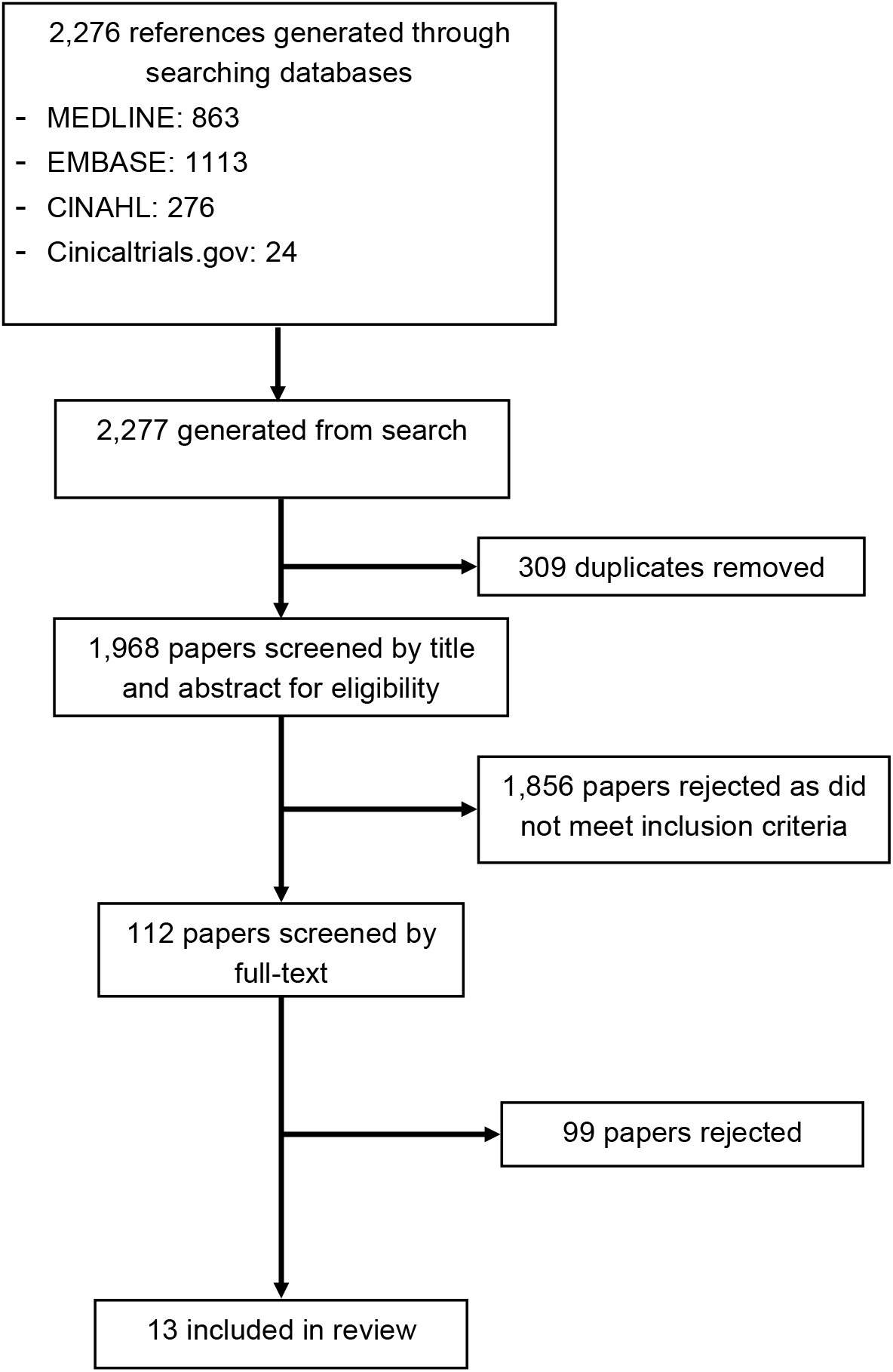
Search Flow Diagram.

### Study Characteristics

Individual study characteristics are presented in *Table 1*.

Twelve studies were conducted in African countries, the other in Iran^13^. Two were in urban areas; ten in rural settings; one was set in both an urban and rural area.

Sample sizes ranged from 81 to 2,712. Enrollment ages differed (*Table 1*), though all children were between 6-60 months. As per inclusion criteria, all studies defined MAM with either WHZ based on *WHO 2006 Growth Standards* and/or MUAC.

All included studies were randomised-controlled trials (RCTs), of which four were cluster-RCTs. The following interventions were compared:

1. Lipid-based nutrient supplements (LNS) versus fortified blended flours (FBF)
2. Comparison of two different formulations of FBF
3. Comparison of two different formulations of LNS
4. A food supplement versus nutrition counselling/usual diet

All products exceeded WHO energy density recommendations (>0·8kcal/g)^14^ (*Appendix 3*).

The primary outcome of all except two studies was the proportion of children with MAM who recovered. The remaining, investigated increments in the fat-free mass index (FFMI) or fat-mass accretion (kg) of participants^15^. One study did not define recovery as obtaining a WHZ > −2, but instead defined recovery as WHZ > −1^13^.

Nevertheless, data was available on the proportions of children who reached a WHZ > −2, which was used in this analysis. The four cluster-RCTs reported outcomes adjusted for the clustered design.

In all studies, once a participant was classified as recovered from MAM, they were discharged from the treatment programme. Two studies treated participants for a maximum of 8 weeks^16^; the remaining trials treated for a maximum of 12 weeks.

### Risk of Bias

*Table 2* summarises the risk of bias assessment. Overall reporting was good, but with numerous areas unclear in several studies. Randomisation methods in *Amegovu 2015*^17^ were rated as having ‘high-risk ‘of bias as allocation of the intervention and control was performed on only two clusters. Furthermore, this study specifically reported that the individual assessing outcomes was not blinded to the intervention.

*Nikiema 2014*^18^ experienced a loss to follow-up rate of 44·6% in the nutrition counselling arm, and therefore was considered to have a ‘high-risk ‘of attrition bias.

Blinding of participants and study personnel was not done in both *Ackatia-Armah 2015*^19^, *Azimi 2020* and *Bailey 2020*^20^, due to clear differences between the two interventions.

## Study Results

### Lipid-Based Nutrient Supplement (LNS) vs Fortified Blended Four (FBF)

Seven trials compared an LNS and a FBF. One study was removed from the final meta-analysis, as it used a type of FBF, corn-soy blend (CSB), which has since been replaced by improved versions (CSB+/++)^16^. The reason was to reduce heterogeneity. Random-effects meta-analysis of the remaining trials (n=7,667) showed that children treated with an LNS were 4% more likely to recover with CSB+/++ (RR 1·05, 95%CI 1·01-1·09, p=0·009) (*Figure 2*). Overall, 42·1% of the variation in relative-risks were attributable to heterogeneity (p=0·125).

**Figure 2:**
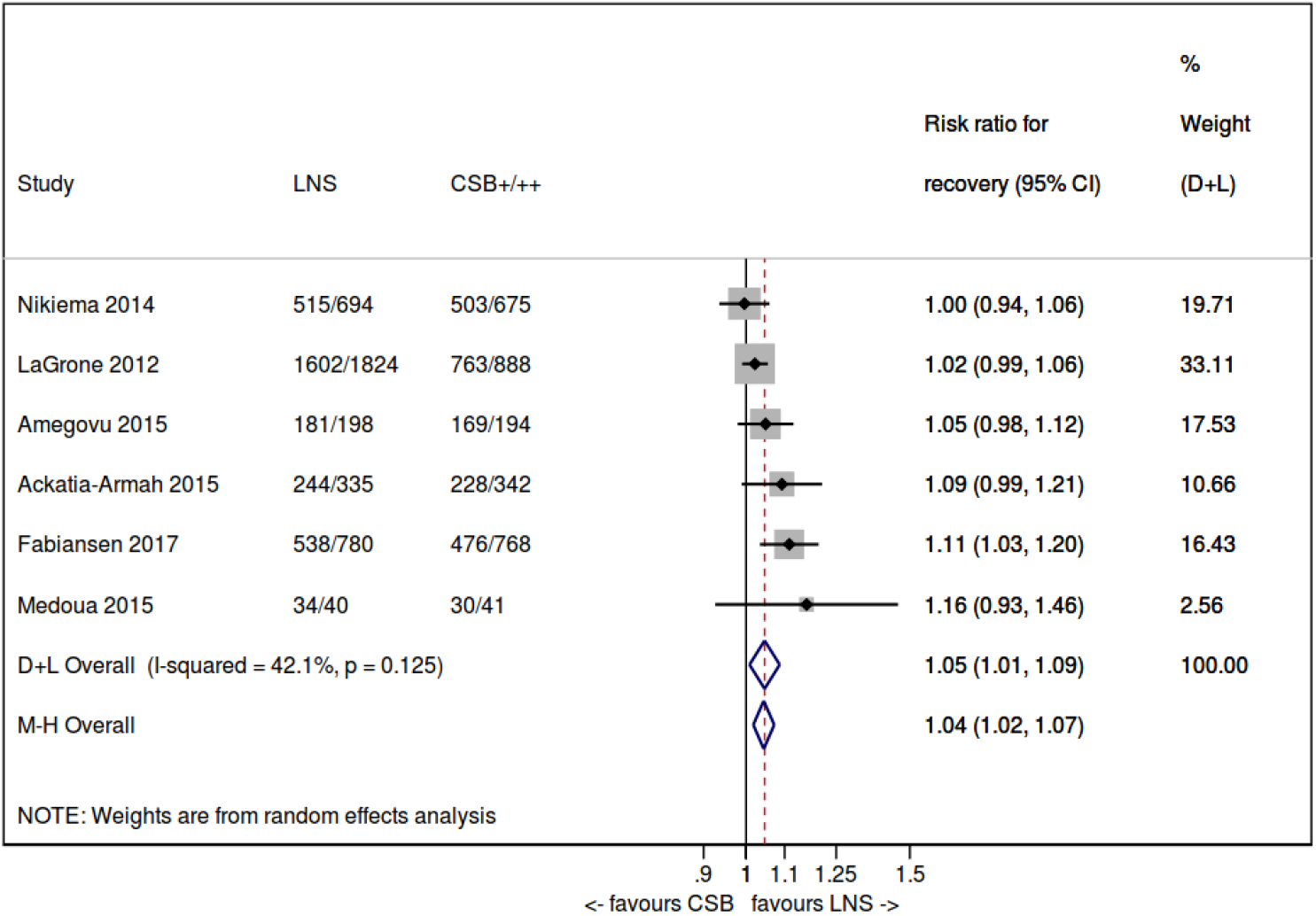
A random-effects meta-analysis of studies comparing the proportions of children who recovered from MAM when treated with LNS vs CSB+/++ Abbreviations: CSB, corn-soy blend; LNS, lipid-based nutrient supplement

A random-effects meta-analysis of four studies (n=5,710) comparing an LNS with CSB+/++, showed that children treated with an LNS have an 18% reduction in the risk of persistent MAM (RR 0·82, 95%CI 0·71-0·95, p=0·009). There was no evidence of heterogeneity across studies (p=0·825).

Progression to SAM whilst receiving either an LNS or a FBF was reported in five trials (n=7,043). Random-effects meta-analysis showed no significant difference in the risk of developing SAM amongst children treated with an LNS compared to FBF (RR=0·87, 95%CI 0·74-1·00, p=0·066). There was no evidence of heterogeneity (p=0·879).

A random-effects meta-analysis of three trials reporting deaths during treatment for MAM with either an LNS or a FBF (n=5,414) showed no differences (RR 0·88, 95%CI 0·47-1·64, p=0·687). There was also no evidence of heterogeneity across studies (p=0·565).

Data from the three trials reporting defaulting from MAM treatment with either an LNS or FBF (n=5,414) showed no evidence of a difference in default rates (RR 1·35, 95%CI 0·96-1·90, p=0·082). There was no evidence of heterogeneity across trials (p=0·307).

One study reported no evidence of a difference in the risk of diarrhoea (p=0·980) or vomiting (p=0·220) during treatment with an LNS versus a FBF^17^.

One study investigated fat-free mass accretion after treatment^15^. This showed that children treated with LNS had, on average, 0·083 kg/m^2^ higher FFMI than those treated with CSB (95%CI 0·003-0·163). Furthermore, there was no evidence of effect modification for the differences in FFMI by season, admission criteria, baseline FFMI, stunting, inflammation, and breastfeeding.

### Different Formulations of LNS

Seven trials compared two different formulations of LNS^15,16,20,21,22,23,24^. Detailed compositions of these LNSs is in *Appendix 3*.

A random-effects meta-analysis of the three trials comparing whey/milk LNS vs soy/no animal product LNS (n=4,948) showed no differences in: recovery (RR 1·01, 95%CI 0·98-1·05, p=0·418, heterogeneity p=0·186); risk of remaining moderately malnourished (RR 1·02, 95%CI 0·77-1·36, p=0·884, heterogeneity p=0·515); risk of progression to SAM (RR 0·95, 95% CI 0·80-1·13, p=0·559, heterogeneity p=0·552); risk of death (RR 0·80 95%CI 0·38-1·66, p=0·542, heterogeneity p=0·676).

Treatment of MAM with a milk-/whey-based LNS was associated with a 39% lower risk of defaulting comparing to treatment with a soy-LNS (n=4,948, RR 0·61, 95%CI 0·40-0·93, p=0·022) (*Figure 3*). There was no evidence of heterogeneity across trials.

**Figure 3:**
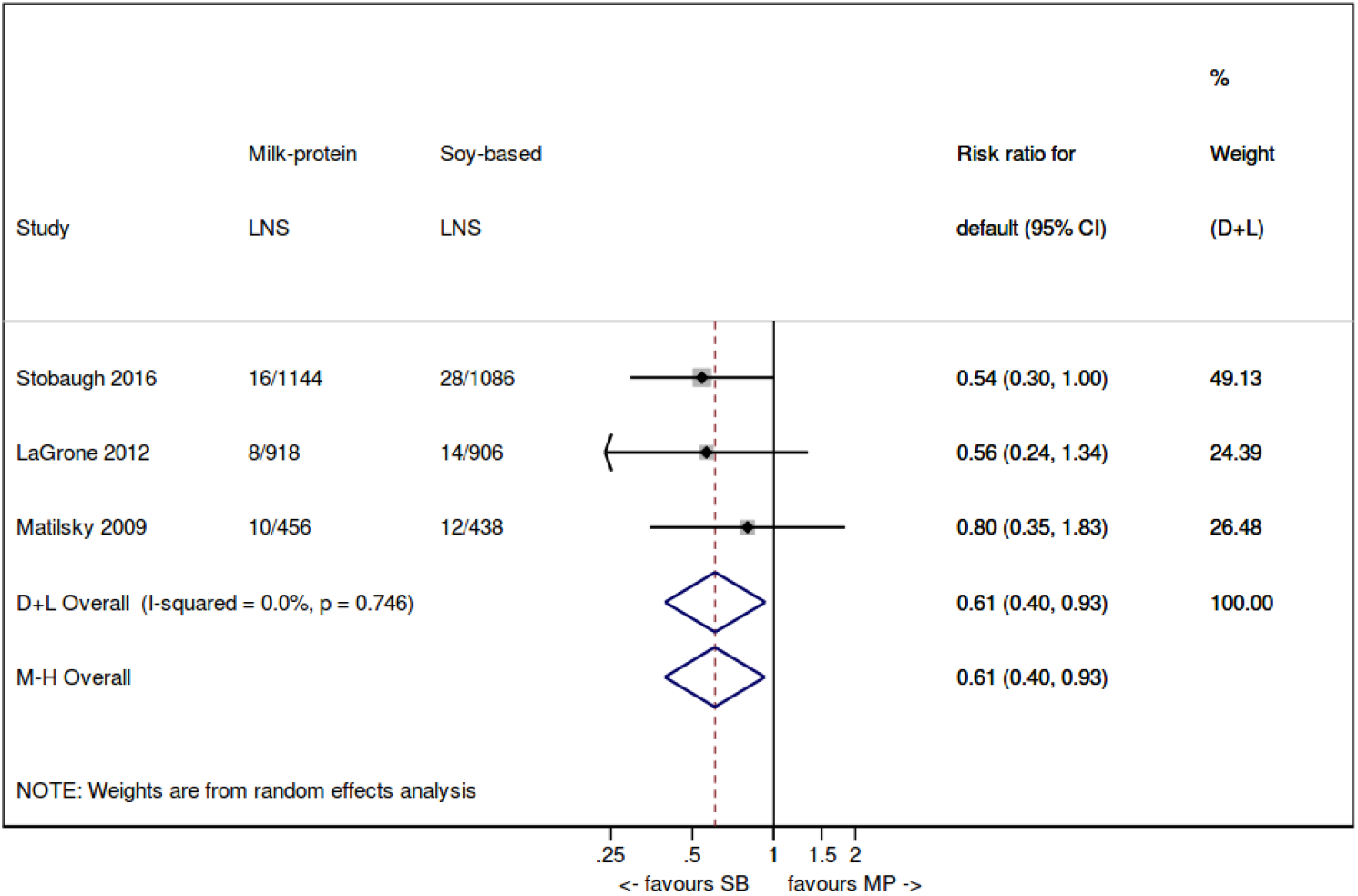
A random effects meta-analysis of studies comparing the relative risk of defaulting in children treated with milk-protein LNS vs soy-based LNS Abbreviations: LNS, lipid-based nutrient supplement; MP, milk-protein; SB, soy-based

A combined treatment of SAM and MAM was investigated in one study: children with MAM were given one sachet of ready-to-use therapeutic food (RUTF) in the combined protocol, and one sachet of ready-to-use supplementary food (RUSF) in the standard protocol. Treatment of MAM with the combined protocol was non-inferior to treatment with standard care (n=1,903, RR 0·00, 95%CI −0·07; 0·07, p=0·97)^20^. At four months post-treatment, there was no significant differences in episodes of diarrhoea (adjusted difference: 0·05, 95%CI −0·9; 1·0, p=0·91), vomiting (adjusted difference: 0·21, 95%CI −1·4; 1·8, p=0·80), fever (adjusted difference: 0·34, 95%CI −0·9; 1·6, p=0·59), cough (adjusted difference: −0·41, 95%CI −1·7; 0·9, p=0·53), and hospitalisation (adjusted difference: 0·52, 95%CI −1·8; 2·9, p=0·66)^25^.

Changes in body composition when treating MAM with two different LNS products was investigated in two studies^15^. One observed that products containing 20% milk protein were better at promoting fat-free mass accretion than products which did not contain milk protein (FFMI difference: 0·097 kg/m^2^, 95%CI −0·002; 0·196). There was no evidence of an effect on FFMI when treating MAM with products that contained 50% milk protein compared to no milk protein (FFMI difference: 0·049 kg/m^2^, 95%CI −0·047; 0·146). At four month follow-up post MAM treatment, no significant differences were observed in fat-free mass accretion when treating with RUTF versus RUSF (adjusted difference: −0·10 kg, 95%CI −0·31; 0·11, p=0·37)^25^.

When considering protein quality, one trial noted no association between LNS protein quality and recovery from MAM (*p* = 0·61)^24^.

In one study, treating MAM using a locally produced LNS (A-RUTF), which substituted half the amount of peanut in standard LNS (S-RUTF) with local soybean and sorghum flours, was associated with significantly lower proportions of children recovering : LNS (A-RUTF: 386 (87·1), S-RUTF: 398 (93·4), p=0·003)^23^. One reason for this was thought to be higher defaulting amongst children supplemented with locally produced LNS (A-RUTF: 56 (12·6), S-RUTF: 27 (6·3), p=0·002).

### Different Formulations of FBF

One study investigated treating of MAM with two different formulations of a FBF; CSB+ and a nutrient-dense malted sorghum-based porridge (MSBP) in Uganda^26^. MSBP substituted a portion of maize in the commercially produced CSB+ with malted sorghum. Recovery from MAM did not differ significantly in children treated with MSBP versus CSB+ (n=94 (90%) vs n=81 (81%), p=0·055).

### Food Supplement vs Nutrition Education / Counselling

Two studies compared treatment of MAM with a food supplement versus a non-food supplement^13,18^. One compared child-centered counselling (CCC) with a blended food (CSB++) and LNS (RUSF) in Burkina Faso; the other compared an LNS (RUSF) with usual diet and nutrition education in Iran.

Children treated with a food product had an 18% increased probability of recovery versus children treated without a supplement (*Figure 4*) (RR 1·16, 95%CI 1·02-1·31, p=0·045). There was evidence of heterogeneity across studies (p=0·045).

**Figure 4:**
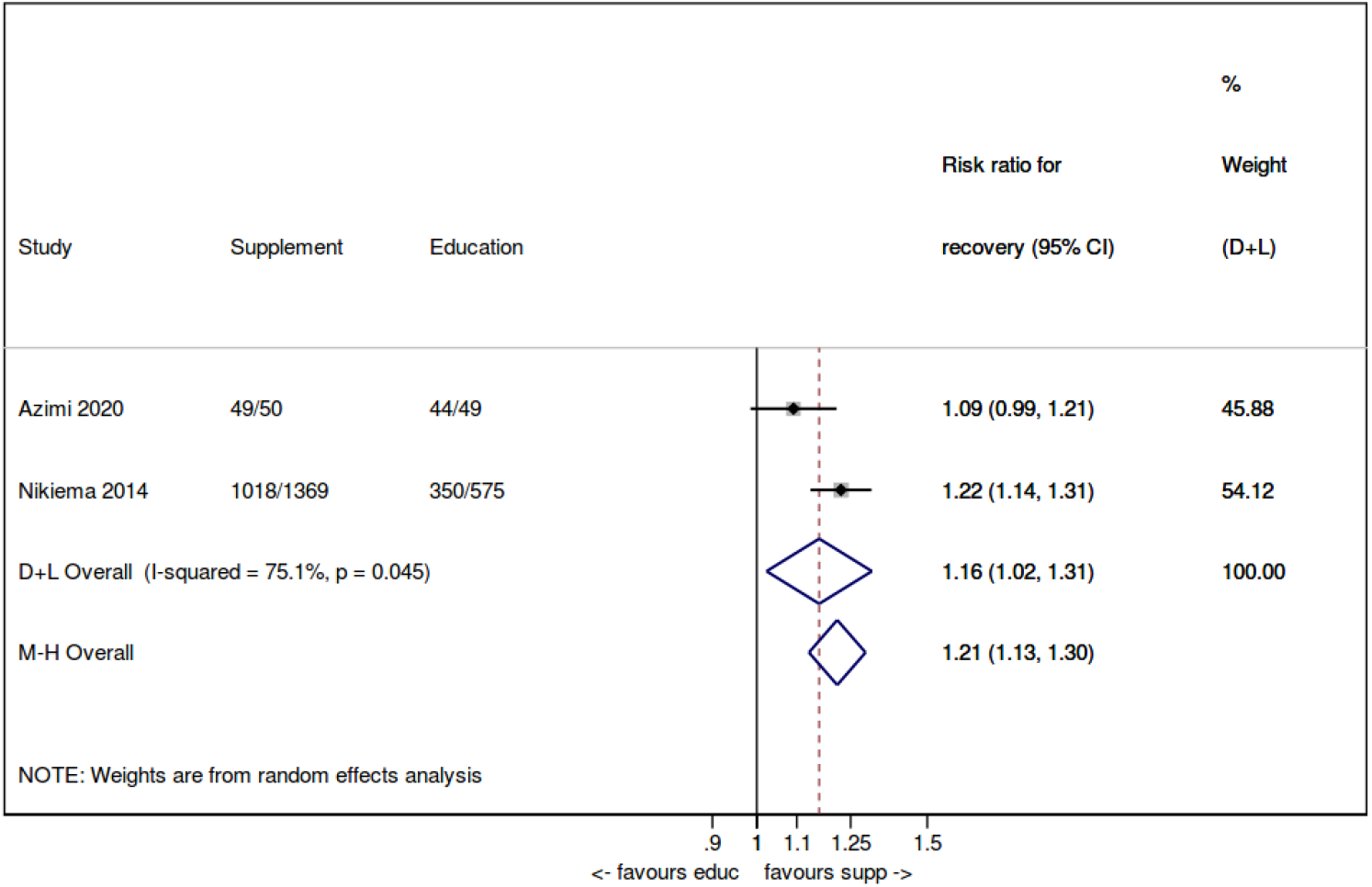
A random effects meta-analysis comparing the probability of recovery in children with MAM treated with a food supplement vs nutrition education alone

In *Azimi 2020* no children progressed to SAM or died and only one child defaulted.

In *Nikiema 2014* children treated with a food product had a 22% reduced risk of progressing to SAM compared to those treated with CCC (RR 0·78, 95%CI 0·62-0·99, p=0·037). There was also strong evidence that children treated with a food product had a 71% reduced risk of defaulting compared to children treated with CCC (RR 0·29, 95%CI 0·17-0·48, p<0·001). Restriction of the analysis to non-defaulters noted the following proportions of children who recovered in each intervention arm: 71·0% recovered with CCC; 77·6% recovered using CSB++ arm; 79·6% recovered using RUSF^18^.

There was no evidence for a difference in the proportions of children who remained moderately malnourished (RR 0·97, 95% CI 0·78-1·21, p=0·786), or died (RR 0·44, 95%CI 0·17-1·17, p=0·101) when treated for MAM with either a food product or CCC.

A significantly lower prevalence of both diarrhoea (n=6 (12%) vs n=14 (28·6%), p=0·01), and fever (8 (16%) vs 18 (36·7%), p=0·05) was observed in children treated with an LNS compared to usual diet^13^.

## Discussion

This was a systematic review and meta-analysis investigating the effectiveness of treatments for moderate acute malnutrition (MAM) in children aged 6-59 months in LMICs. Ten of the thirteen included trials compared the effectiveness of different supplementary food products in promoting MAM recovery, reflecting the recent focus on the use of products in MAM management^6^. Children treated with an LNS, had a 5% increased probability of recovery compared to those treated with CSB++. Furthermore, treatment with an LNS was associated with a lower risk of persistent MAM, compared with a FBF. Treatment with a food product was associated with a higher probability of recovery, a lower risk of progressing to SAM, and a lower risk of defaulting, compared to children who did not receive supplementation.

In the past, others have also reviewed MAM management^27,28,29,30,31^. Results for our primary outcome – recovery – are consistent with others’ findings that treating MAM with an LNS was associated with an increased relative risk of recovery, compared to treatment with FBFs: *Lazzerini 2013* (RR 1·04, 95%CI 0·99-1·09), *Lenters 2013* (RR 1·11, 95%CI 1·04-1·18), *Gera 2017* (RR 1·08, 95%CI 1·02-1·14), and *Das 2020* (RR 1·07, 95%CI 1·02-1·13). All reported relatively few studies, often with significant heterogeneity between studies even after attempts at stratification and subset analysis. The major step forward we took was to only include papers defining MAM using the *WHO 2006 Growth Standards (*rather than the old *NCHS References*. This affects the profile of children enrolled and explains greater homogeneity in our results overall^32^. Since *WHO 2006 Growth Standards* are now common worldwide, our approach also better represents the current population of interest and our findings enable more generalisable conclusions to be drawn. Another key benefit of our review is to present latest research in this area – hence critical to the upcoming 2021 WHO guidelines process^7^.

The following may contribute to the lower recovery rates seen amongst MAM children treated with FBFs compared with LNSs: 1) FBFs resemble staple foods and thus may be more openly shared amongst other household members^33,34,35^. 2) Children must consume roughly eight-times the mass of FBF compared to LNS, thus potentially discouraging breastfeeding / eating other foods provided at home^33,35^. 3) FBFs are lower in fat content and energy density compared to LNSs (*Appendix 2*). This is supported by evidence showing that LNS supplementation does not replace other foods in the diet^36,37^, is highly acceptable in an African context^38,39^, and is shared amongst household members substantially less than FBFs^40,41^.

This review has highlighted some key gaps in the current body of evidence, which can be grouped into population, intervention/comparator and outcome related.

Out of thirteen included studies, twelve were conducted in African populations, making it difficult to generalise conclusions to South Asian contexts. This is problematic given that over half of all wasted children globally live in South Asia^1^, and likely have different growth trajectories and energy needs compared to children from Africa^20^. Though promising early evidence highlights the effectiveness and acceptability of LNSs in South Asia^42^, more research is needed to understand similarities and differences compared with other populations.

Another striking feature of studies in our review is that all used active controls. This risks underestimating the true benefit of MAM treatment since the field reality is that children with MAM are neither identified not get any treatment in many settings worldwide. Future studies should consider more ‘usual care’ controls (i.e. minimal/no specific care) to better understand true impact of MAM care. This would be ethically acceptable in some settings in light of a recent recommendation that children with MAM presenting to primary care should not be routinely treated^43^. Cost-effectiveness data would be another important part of such work.

In eleven included trials, anthropometry and in particular recovery of ‘normal’ weight was the main outcome measure. Whilst anthropometry is widely used as the key measure of nutritional status, it is malnutrition-associated risk of mortality and morbidity that really matters rather than body size alone^44^. Wasting, low WHZ, which defines MAM is strongly associated with high risk of mortality^45,46^. However, there is increasing evidence that weight recovery does not necessarily mean return to low clinical risk of those who were never malnourished: children who have had MAM remain at high risk of relapse post treatment^47^. Children with MAM, who recovered as part of the *LaGrone 2012* study, were followed-up for a further 12-month period in order to assess the long-term effects of treatment with CSB++ and RUSF^48^. During this period, only 63% of children remained well-nourished, highlighting the vulnerability of children post MAM recovery according to anthropometric measures alone. Measuring the proportions of children who progress to SAM, require admission or die during treatment, are more meaningful measurements of short-term clinical effectiveness of interventions, as these highlights unwanted negative outcomes along the spectrum of anthropometric status. However, these outcomes are relatively rare, can be more complex to measure and certainly in the case of mortality require far sample sizes for robust analysis – so are not commonly included in research.

Other more clinically meaningful outcome measures include body composition and it is encouraging to see studies in our review starting to assess this^49^. Body composition is especially valuable since it also gives information about potential longer term future risk, including of NCD^50^. There is concern that routine supplementation of MAM children with energy-dense products will encourage unhealthy weight-gain that risks predisposition to NCDs^51^. Balancing this evidence from one of the studies in our review suggests the opposite; that the majority of weight gain during supplementation with an energy-dense product is fat-free mass^15^, and that there are no significant differences in changes in body composition depending on the energy density of products^25^. Both these findings persist up to four months post treatment of MAM^25^.

Another important consideration in the treatment of MAM, is its relation to SAM treatment. Although both conditions lie on a continuum of acute malnutrition, both are managed in separate programmes, using different food products and under two organisations; UNICEF for SAM and WFP for MAM. A cluster-RCT based in Kenya and South Sudan investigated a combined protocol for treating acute malnutrition compared to standard care^20^. They noted no significant difference between recovery rates (risk difference 0·03, 95%CI −0·05-0·10, p=0·52), and cost-effectiveness of the combined protocol (US$123 less per child recovered) compared to standard care. These results are echoed in *Maust 2015*^52^. Overall, combined management may be simpler, more cost-effective and reach more children.

We acknowledge some limitations of our review and meta-analysis: 1) Relevant papers could have been missed by restricting the search to papers published after 2006 and limiting to English language. 2) In the risk of bias assessment, we did not search for original paper protocols when determining selective reporting, and therefore, it is unclear as to how much reporting bias contributed to concealment of undesirable results. This was not considered an issue for the primary outcome, recovery, as all papers provided relevant data. 3) We only found a small number of eligible papers, and thus generalisability may be limited, particularly as most papers were conducted in African LMICs. Nevertheless, the small quantity of papers identified highlights the need for more robust research in MAM management, specifically those that address points raised in this review.

## Conclusion

How best to treat children with MAM is a key question for global child health. We found most current evidence focuses on the use of food supplements and involves studies with active controls rather than no-treatment control as is often the case in everyday practice. This risks underestimating the true benefits of treatment. In studies we identified, supplementation with LNS improves anthropometric recovery and prevents progression to SAM compared to supplementation with FBF. The role of nutrition counselling/education alone, or in combination with a supplementary food-product, warrants further research, particularly in areas with good food security. Outcomes in current research focus on weight-recovery: future trials should include more clinically-meaningful outcomes, such as progression to SAM, admission, death and/or changes in body composition.

|  |  |
| --- | --- |
| **Author Contributions** | Gluning, I – designed the search strategy, conducted the search and meta-analysis, and wrote the initial manuscript.<br>Kerac M – conceived the project, supervised and provided feedback on manuscript drafts.<br>Bander A – conducted a second search and review of references<br>Bailey J – provided feedback on manuscript drafts.<br>Opondo C – conceived the project, supervised, assisted in meta-analysis and provided feedback on drafts of manuscript<br>All authors: read and approve of final version of the manuscript |
| **Declaration of Interest** | The authors have declared that no competing interests exist. |
| **Funding** | There was no funding source. |

## Supporting information

MAM Review Appendix

MAM Review PRISMA Checklist

## Data Availability

Systematic review of the literature.
Meta-analysis pooled data and do-files available upon request to corresponding author.

## Abbreviations

AM: Acute Malnutrition
CCC: Child Centred Counselling
CCT: Controlled-Clinical Trials
CI: Confidence Interval
ComPAS: Combined Protocol for Acute Malnutrition Study
CSB: Corn-Soy Blend
FBF: Fortified Blended Flours
FFM: Fat-Free Mass
FFMI: Fat-Free Mass Index
HAZ: Height-for-Age z-score
HFIAS: Household Food Insecurity Access Scale
HIV: Human Immunodeficiency Virus
LMIC: Low- and Middle-Income Country
LNS: Lipid-based Nutrient Supplement
MAM: Moderate Acute Malnutrition
MSBP: Malted-Sorghum Based Porridge
MUAC: Middle Upper Arm Circumference
NCHS: National Centre for Health Statistics
PICO: Point, Intervention, Comparator, Outcome
PRISMA: Preferred Reporting-Items for Systematic Reviews and Meta-Analyses
RCT: Randomised-Controlled Trial
RR: Relative Risk
RUF: Ready-to-Use Food
RUSF: Ready-to-Use Supplementary Food
RUTF: Ready-to-Use Therapeutic Food
SAM: Severe Acute Malnutrition
SFP: Supplementary Feeding Programme
TB: Tuberculosis
UN: United Nations
UNICEF: United Nations Children’s Fund
WFP: World Food Programme
WHO: World Health Organisation
WLZ: Weight-for-length z-score
WHZ: Weight-for-Height z-score

