## Supplementary material for "The management of Moderate Acute Malnutrition (MAM) in children aged 6-59 months: A systematic review and meta-analysis": MAM Review Appendix

### *Appendix 1 – Search Strategy for MEDLINE and EMBASE*

1. exp Malnutrition/
2. exp Protein-Energy Malnutrition/
3. exp Child Nutrition Disorders/
4. exp Wasting Syndrome/
5. “moderate acute malnutrition”.mp.
6. (moderate* adj2 malnutrition).mp.
7. (moderate* adj2 malnourish*).mp.
8. (moderate* adj2 undernutrition).mp.
9. (moderate* adj2 wast*).mp.
10. “mid* upper arm circumference”.mp.
11. (mid* upper arm circumference adj2 “125”).mp.
12. “MUAC”.mp.
13. *1 OR 2 OR 3 OR 4 OR 5 OR 6 OR 7 OR 8 OR 9 OR 10 OR 11 OR 12*
14. exp Infant/
15. exp Child/
16. exp Child, Preschool/
17. *14 OR 15 OR 16*
18. exp Dietary Supplements/
19. exp Food, Fortified/
20. exp Food, Formulated/
21. ((fortif* or enrich* or formulat* or supplement* or lipid* or blend*) adj2 (food* or spread* or diet* or feed* or product*)).mp.
22. “ready to use food*”.mp.
23. “ready to use supplementary food”.mp.
24. “RUSF”.mp.
25. “lipid-based nutrient supplement”.mp.
26. (lipid-based adj2 supplement*).mp.
27. “LNS”.mp.
28. Nutributter.mp.
29. “ready-to-use therapeutic food”.mp.

30. “RUTF”.mp.

31. (therapeutic adj2 food).mp.

32. plumpy*.mp.

33. “FBF”.mp.

34. “fortified blended flour”.mp.

35. super-cereal*.mp.

36. “corn soy* blend*”.mp.

37. “wheat soy* blend*”.mp.

38. “rice milk blend*”.mp.

39. “milk rice blend*”.mp.

40. “pea wheat blend*”.mp.

41. “cereal pulse blend*”.mp.

42. “CSB”.mp.

43. “CSB+”.mp.

44. *18 OR 19 OR 20 OR 21 OR 22 OR 23 OR 24 OR 25 OR 26 OR 27 OR 28 OR 29 OR 30 OR 31 OR 32 OR 33 OR 34 OR 35 OR 36 OR 37 OR 38 OR 39 OR 40 OR 41 OR 42 OR 43*

45. exp Education/

46. exp Health Education/

47. exp Counseling/

48. “standard care”.mp.

49. “basic care”.mp.

50. “control”.mp.

51. (education adj2 nutrition).mp.

52. (counseling adj2 nutrition).mp.

53. ((local or home) adj2 (food* or feed* or diet*)).mp.

54. *45 OR 46 OR 47 OR 48 OR 49 OR 50 OR 51 OR 52 OR 53*

55. *13 AND 17 AND 44 AND 54*

56. Limit 55 to (English language and humans and yr=”2006-Current”)

*Appendix 2 – Reasons for exclusion following screening of full-text*

| Study | Reason for Exclusion |
|---|---|
| **Aboud 2008** | Enrolled children did not have MAM |
| **Aburto 2010** | Investigated effect of micronutrient supplementation on physical activity |
| **Ackatia-Armah 2012** | Full text not available |
| **Addo 2020** | Investigated improvements in gross motor and communication scores |
| **Ahmed 2019** | Study protocol for ongoing trial |
| **Akhter 2011** | Preventative management of undernutrition |
| **Amagloh 2012** | Investigated composition of sweet potato based complementary food |
| **Annan 2014** | Technical brief |
| **Arimond 2015** | Preventative management of undernutrition |
| **Ashton 2011** | Preventative supplementation |
| **Ashworth 2009** | Investigated the likelihood of effect of nutrition education programmes |
| **Batra 2016** | Enrolled children did not have MAM |
| **Bazzano 2017** | Literature review |
| **Borg 2017** | Enrolled children did not have MAM |
| **Borg 2019** | Lessons learned when testing a novel supplementary food |
| **Borg 2020** | Preventative supplementation |
| **Briend 2011** | Population based mathematical model |
| **Brown 2009** | Literature review |
| **Chanani 2019** | No comparator treatment group. |
| **Chang 2013** | Follow-up study post treatment of MAM |
| **Choudhury 2016** | Defined undernutrition using weight-for-age |
| **Cohuet 2012** | Investigated acceptability and intra-household use of supplements |
| **Daniel 2019** | Investigated dietician impact in hospital setting |
| **Das 2019** | Literature review |
| **Das 2020** | Systematic review |

| | |
|---|---|
| **Delchevalerie 2015** | Defined MAM using Department for Health and Human Statistics |
| **Desai 2014** | Retrospective cohort study |
| **Dido 2011** | Full text not available |
| **Egata 2014** | Investigated predictors of acute undernutrition |
| **Fatima 2018** | Enrolled mildly malnourished children (WHZ>-2 to <-1) |
| **Fernald 2016** | Investigated impact of supplementation on stunting |
| **Fitriyanto 2019** | No comparator treatment group. |
| **Flax 2013** | Investigated feeding behaviours with supplementation |
| **Flax 2015** | Investigated dietary intake during supplementation |
| **Friis 2015** | Literature review |
| **Gaffey 2013** | Investigated management of moderate diarrhoea |
| **Gigante 2007** | Enrolled school-aged children |
| **Hossain 2012** | Conference proceeding - full text unavailable |
| **Huybregts 2012** | Preventative management of undernutrition. Undernutrition defined using NCHS references. |
| **Huybregts 2017** | Preventative management of undernutrition |
| **Iannotti 2015** | Enrolled children aged 3-13 years. |
| **Ickes 2015** | Diet adequacy assessment |
| **Ikawati 2019** | Defined undernutrition using weight-for-age |
| **Inayati 2012** | Enrolled children did not have MAM - had mild wasting (defined as a WHZ>-1.5 to <-1 |
| **Isanaka 2010** | Preventative supplementation |
| **Iuel-Brockdorf 2017** | Investigated home behaviours during supplementation |
| **James 2016** | Prospective cohort study |
| **Juliana 2017** | Investigated zinc deficiency |
| **Kajjura 2019** | Investigated hygiene practice and knowledge of mothers following nutrition education |
| **Karakochuk 2015** | Defined MAM using NCHS references |
| **Kekalih 2019** | Study protocol for ongoing trial |
| **Khan 2013** | No comparator treatment group. Enrolled children aged 6 months to 8 years |
| **Khatib 2010** | Descriptive study of a vulnerable group |
| **Kimani-Murage 2013** | Full text not available - enrolment of well nourished mother-child pairs |

| | |
|---|---|
| **Kulwa 2014** | Study protocol for ongoing trial |
| **Langendorg 2014** | Preventative supplementation |
| **Lanou 2019** | Enrolled children did not have MAM |
| **Lazzerini 2013** | Systematic review |
| **Lelijveld 2020** | Follow-up study 4 months post treatment of MAM |
| **Linneman 2007** | No comparator treatment group. |
| **Magnin 2017** | No comparator treatment group. |
| **Maheswari 2012** | Used weight to define undernutrition |
| **Manary 2020** | Literature review |
| **Mangani 2014** | Investigated morbidity in well nourished children |
| **Marron 2015** | Full text not available |
| **Matondo 2016** | Defined MAM using NCHS references |
| **Maust 2015** | Combined protocol for MAM and SAM treatment. Results published do not differentiate between MAM and SAM |
| **Moramarco 2016** | Retrospective observational study |
| **Moramarco 2018** | Retrospective observational study |
| **Mostafa 2020** | Proof-of-concept study - full text not available - study not complete |
| **Nackers 2010** | Defined MAM using NCHS references |
| **Nane 2019** | Study protocol for ongoing trial |
| **Olney 2017** | Preventative supplementation - primary outcome assessed anaemia |
| **Pereira 2020** | Investigated glucose levels and Hb with supplementation |
| **Perez-Exposito 2009** | Literature review |
| **Phuka 2009** | Measured mean anthropometric changes. Primary outcome was reduction in stunting. |
| **Phuka 2011** | Investigated acceptability of LNS in Malawi - amount consumed, time to eat and maternal rating |
| **Prado 2016** | Enrolled children did not have MAM |
| **Puett 2013** | Cost-effectiveness analysis |
| **Pulakka 2017** | Enrolled children did not have MAM - measured impact of supplementation on physical activity |
| **Ragini 2014** | Primary outcome focused on mothers' continuation of nutrition education practices |
| **Rogers 2017** | Behaviour change and cost-effectiveness analysis |

| | |
|---|---|
| **Ruel 2008** | Compared preventative supplementation with recuperative |
| **Sachdeva 2014** | Severe acute malnutrition |
| **Saleem 2014** | Enrolled children did not have MAM |
| **Sayyad-Neerkorn 2015** | Preventative supplementation |
| **Scherbaum 2015** | Recovery defined as WHZ>-1.5 |
| **Schlossman 2015** | Defined undernutrition using weight-for-age and height-for-age |
| **Schlossman 2017** | Defined undernutrition using weight-for-age and height-for-age |
| **Schlossman 2018** | Enrolled school-aged children |
| **Segre 2017** | Investigated economic impacts of local vs offshore therapeutic supplements |
| **Sguassero 2012** | Preventative supplementation |
| **Shen 2017** | Cost-effectiveness analysis |
| **Siega-Riz 2014** | Supplementation effect on micronutrient status |
| **Somasse 2013** | Follow-up study post treatment of MAM |
| **Somasse 2016** | Management of severe acute malnutrition |
| **Steenkamp 2015** | No comparator treatment group. |
| **Stobaugh 2017** | Investigated children post recovery from MAM |
| **Stobaugh 2017** | Investigating sustaining recovery post treatment of MAM |
| **Stobaugh 2018** | Investigated factors associated with relapse post MAM treatment |
| **Tandon 2018** | Investigated effect of supplementation on neutropenia in cancer patients with acute malnutrition |
| **Taye 2016** | Descriptive study |
| **Thakwalakwa 2015** | Defined MAM using NCHS references |
| **Trehan 2015** | Investigated an extended course of treatment for MAM post recovery |
| **Van der Kam 2016** | Preventative supplementation |
| **Van Waardenburg 2009** | Investigated critically ill infants with bronchiolitis |
| **Verna 2012** | Conference proceeding - full text unavailable. Did not use WHO growth standards to define MAM |
| **Vray 2018** | Study protocol for ongoing trial |
| **Wang 2013** | Investigated acceptability and feeding practices in MAM treatment |

| | |
|---|---|
| **Wilner 2017** | Lessons learned / communication whilst implementing a supplementary feeding programme |
| **Zhang 2019** | Systematic review |

*Appendix 3 – Nutrient compositions of interventions by study and including WHO Recommendations for children aged 6-59 months*

| Study | Intervention | Composition | | | | |
|---|---|---|---|---|---|---|
| | | Dry mass of food (g) | Energy (kcal) | Energy Density (kcal/g) | Protein (g) | Fat (g) |
| ***WHO Recommendations*** | per 1000kcal of Supplement (70% of daily energy intake) | - | 1000 | 0.8 | 20-43 | 25-65 |
| ***Matilsky 2009*** | CSB | 200 | 749 | 3.7 | 34 | - |
| | Milk/peanut FS & Soy/peanut FS | 136 | 749 | 5.5 | 19 | - |
| ***LaGrone 2012*** | CSB+ | 143 | 563 | 3.9 | 21 | 13 |
| | Soy RUSF | 104 | 563 | 5.4 | 17 | 40 |
| | Soy/whey RUSF | 103 | 563 | 5.5 | 15 | 38 |
| ***Nikiema 2014*** | CSB++ | 65 | 273 | 4.2 | 10 | 6 |
| | RUSF | 50 | 258 | 5.2 | 9 | 17 |
| ***Medoua 2015*** | CSB+, oil | 72 | 320 | 5 | 10 | 11 |
| | RUSF | 61 | 320 | 5.2 | 10 | 21 |
| ***Amegovu 2015*** | CSB+ | 269 | 1,200 | 4 | 46 | 30 |
| | SPB | 269 | 1,228 | 5 | 40 | 48 |
| ***Ackatia-Armah 2015*** | RUSF | 92 | 500 | 5.4 | 13 | 33 |
| | CSB++ | 127 | 501 | 3.9 | 18.4 | 12 |
| | Misola | 125 | 500 | 4 | 18.5 | 14 |
| | LMF + MNP | 129 | 500 | 3.9 | 15.5 | 14 |
| ***Stobaugh 2016*** | Soy RUSF | 105 | 560 | 5.3 | 17 | 37 |
| | Whey RUSF | 105 | 516 | 4.9 | 11 | 36 |
| ***Kajjura 2019*** | MSBP | 150 | 675 | 4.5 | 26 | 6.6 |
| | CSB+ | 158 | 600 | 3.8 | 25 | 9.5 |
| ***Kohlmann 2019*** | A-RUTF | 100 | 560 | 5.6 | 14.5 | 29.2 |
| | S-RUTF | 100 | 559 | 5.6 | 15.8 | 33 |

| | | | | | | |
|---|---|---|---|---|---|---|
| ***Azimi 2020*** | RUSF | 95 | 475 | 5 | 15 | 23 |
| ***Roediger 2020*** | HiPro-RUSF | 100 | 530 | 5.3 | 13.5 | 32.8 |
| | C-RUSF | 100 | 537 | 5.4 | 13.4 | 32.9 |
| ***Bailey 2020*** | RUSF | - | 500 | - | - | - |
| | RUTF | - | 500 | - | - | - |
